# Trajectories of Cognitive Decline Before and After New-onset Hypertension

**DOI:** 10.1101/2024.08.03.24311456

**Authors:** Qingmei Chen, Jianye Dong, GC Chen, Haibin Li, Yueping Shen, Jianian Hua

**Affiliations:** Department of Physical Medicine &Rehabilitation, The First Affiliated Hospital of Soochow University, Suzhou, 215000, Jiangsu Province, China; Department of Obstetrics and Gynecology, The First Affiliated Hospital of Soochow University, Suzhou, Jiangsu, China; Department of Nutrition and Food Hygiene, Suzhou Medical College of Soochow University, 199 Ren’ai Road, Suzhou, 215123, China; Department of Cardiac Surgery, Heart Center and Beijing Key Laboratory of Hypertension, Beijing Chaoyang Hospital, Capital Medical University, Beijing, China; Department of Epidemiology and Biostatistics, School of Public Health, Medical College of Soochow University, 199 Ren’ai Road, Suzhou, 215123, China; Department of Neurology, The First Affiliated Hospital of Soochow University, Suzhou, Jiangsu, China

**Keywords:** hypertension, cognition, cognitive decline, aging, cohort study, UK population

## Abstract

**Background:** Hypertension is a known factor for cognitive impairment, especially in midlife. However, whether the cognitive function declines before and shortly after new-onset hypertension remains largely unknown.

**Objectives:** We aimed to examine the cognitive trajectories before and after new-onset hypertension among community-dwelling midlife and older participants.

**Methods:** This study included 2,964 participants from the English Longitudinal Study of Ageing who were free of hypertension at baseline. Participants who had a stroke at baseline or during follow-up were excluded. Global cognition (a summary of semantic fluency, orientation, and memory) was assessed at baseline (wave 2, 2004) and at least once from wave 3 to wave 9 (2018). New-onset hypertension was defined by self-reported doctor diagnosis, use of antihypertensive medications, and blood measurements < 140/90 mmHg.

**Results:** Over a median follow-up of 13.6 years, 1,121 (37.8%) participants developed hypertension. The cognitive decline rate among those who later developed hypertension during the pre-hypertension period was similar to the rate among those who remained hypertension-free throughout the study. After the onset of hypertension, the rate of cognitive decline accelerated in global cognition (β, −0.015 SD/year; 95% CI, −0.026 to −0.003; *p*=0.011), semantic fluency (β, −0.015 SD/year; 95% CI, −0.027 to −0.003; *p*=0.017), and memory (β, −0.022 SD/year; 95% CI, −0.033 to −0.010; *p*<0.001), but not in orientation ability (β, −0.012 SD/year; 95% CI, −0.028 to 0.005; *p*=0.157). Participants who developed hypertension in older age did not experience a reduced impact of post-hypertension cognitive decline compared to those who developed hypertension in midlife.

**Conclusions:** Participants experienced accelerated cognitive decline upon developing new-onset hypertension. Older participants are equally susceptible to cognitive impairment due to hypertension. Early antihypertensive initiation is crucial in both midlife and later life to protect cognitive health.

## Introduction

More than 60 years ago, Spieth discovered that pilots and air traffic control specialists with untreated hypertension had poorer performance on a battery of psychological tests.(1) A meta-analysis including 136 articles estimated that individuals with hypertension had a 1.19 to 1.55 higher risk of developing cognitive impairment, Alzheimer’s Disease (AD), and dementia.(2) Hypertension affects a billion people worldwide, making it the most prevalent and one of the most modifiable risk factors for dementia prevention.(3) Large-scale meta-analyses published in 2021 and 2023 suggested that the use of antihypertensive agents was associated with a lower risk of incident dementia.(4,5) However, a key challenge is that most studies only focused on participants with prevalent hypertension. Data on new-onset hypertension is scarce.(6) Previous research only had cognitive data after the study baseline, at which time hypertension had existed for several years, lacking cognitive data shortly after hypertension onset or the timeline of cognitive decline. If the cognitive function deteriorates shortly after or even before hypertension onset, then the current management for hypertension is insufficient. Moreover, hypertensive participants are more likely to have metabolic symptoms such as diabetes, dyslipidemia, and obesity. Therefore, we are not sure whether their cognitive function had already declined before hypertension onset.(7) To our best knowledge, no study has estimated the cognitive trajectories before and after new-onset hypertension at the same time.

Prior studies on age difference in the relationship between hypertension and cognitive health have yielded mixed results. For late life (65—84) and the oldest (≥85) hypertension, studies have reported that higher blood pressure (BP) was associated with better cognitive function.(8) A meta-analysis including participants aged ≥65 also suggested that hypertension is a protective factor for dementia.(9) On the contrary, other studies found that hypertension was associated with faster cognitive decline,(10) a U-shaped association,(11) or no association.(12) Unlike late-life hypertension, midlife hypertension is confirmed to be a strong risk factor for dementia.(2,13) The ‘age difference’ has become a hot topic in recent years because increased dementia risks have been repeatedly linked to younger age at the onset of cardiovascular disease, such as diabetes, heart failure, and atrial fibrillation.(14–16) Hence, there is a need to explore the age difference in the association between new-onset hypertension and subsequent cognitive decline.

Using data from the English Longitudinal Study of Ageing (ELSA), a study with repeated cognitive assessments and a follow-up of 16 years, we aimed to compare the trajectories of cognitive decline before and after new-onset hypertension and perform subgroup analysis according to age and other risk factors.

## Methods

### Study population

ELSA is a nationally representative longitudinal study of a community-dwelling English population aged ≥50. A detailed study design is available elsewhere.(17) The wave 1 survey recruited 12,099 participants between 2002 and 2003. Follow-up waves were conducted every two years, with the latest published one being in wave 9 (2018—2019). The ELSA study received approval from the London Multicenter Research Ethics Committee. All participants signed the informed consent.

For this study, we combined data from wave 2 to wave 9 and defined wave 2 as the baseline for the following reasons: a) handling the practice effect of cognitive tests,(18,19) missing blood pressure measurement in wave 1, and higher follow-up rates. At wave 2, 9,269 participants underwent cognitive assessments, and 5,680 of them were excluded due to being less than 50 years old (n=252), history of Parkinson’s disease/dementia/AD (n = 104), stroke (n = 464), or with prevalent hypertension at baseline (n=5,350). Among the remaining 3,589 participants, we removed 143 participants who had an incident stroke during the follow-up, 483 participants who did not receive at least one cognitive test from wave 3 to wave 9 (Supplemental Table 1), and 1 participant for handling missing data. We finally included 2,964 participants free of hypertension at baseline (Figure 1).

**Figure 1.** Flow gram of cohort creation.

### Ascertainment of new-onset hypertension

In each wave, self-reported doctor-diagnosed hypertension and usage of antihypertensive medications were recorded, both of which were considered as indicators of hypertension. We defined the time of hypertension onset as the midpoint between the last interview without hypertension and the first interview reporting hypertension. In waves 2, 4, 6, and 8, participants underwent 0 to 3 BP measurements. BP was measured by the nurse on the right arm of each participant while they were sitting, using the Omron HEM-907 device. The first reading was taken after a 5-minute rest period. We collected the average BP of two or three BP readings. For participants unaware of their hypertension, the time of hypertension onset was defined as the midpoint between the last time BP<140/90 and the first time BP≥140/90.

### Cognitive function

Cognitive function was assessed in each wave. Three domains were analyzed in this study: semantic fluency, orientation, and memory. The semantic fluency test requested participants to name as many animals as possible in one minute. The score ranged from 0 to 50, with a median score (interquartile range) of 21 (17, 25) in wave 2. Orientation ability was assessed by asking the year, month, date, and the day of the week, with scores ranging from 0 to 4. Memory score was evaluated by immediate and delayed recall of ten words. One point for each correct word and the maximum score is 20. The three tests were widely used and well-documented by previous studies.(20,21) We performed z-transformation on the cognition score of each individual (*i*) in each time point (*t*) for comparability of the effect sizes across different cognitive tests, using the formula: z-score_it_ = (*score_it_* - *mean score_baseline_*) / *SD_baseline_*. A *z* score of −1.000 at any time point represented 1.000 SD below the mean cognitive score at baseline. To summarize the results of the three cognitive tests, a global cognition z score was created by averaging the z scores of the three tests and then standardizing it to baseline using the same manner.

### Covariates

We adjusted for covariates associated with hypertension or cognitive function, which included age, sex, education level, marital status, current smoking, alcohol drinking, body mass index (BMI), physical activity, depressive symptoms, diabetes, cancer, chronic lung diseases, and heart failure.(22) Details of covariates and handling of missing covariates are provided in Supplemental Methods 1 and 2, and Supplemental Tables 2 and 3.

### Statistical Analysis

We calculated the standardized mean difference (SMD) before and after the inverse probability of treatment weight (IPTW) to assess the balance between the hypertension-free group and the hypertension group in which participants developed hypertension between waves 2 to 9. Our primary analysis was a linear mixed-effect model, including both fixed effects and random effects. This model allowed for measurements of cognitive test scores at flexible time points. We fitted fixed effects for intercept, slopes, and all covariates, and random effects for intercept and slopes to account for repeated scores on the same participant, variations in baseline scores, and variations in slopes across participants. A detailed description of this model is shown in Supplemental Figure 1. Figure 2 illustrates the cognitive trajectories of the two groups, comprising the following kinds of trajectories: the average decline rate from baseline to the end of the follow-up of the entire hypertension-free group, and the average difference in 1) baseline scores between the two groups, 2) slope during the whole follow-up period for the hypertension-free group vs. slope of the hypertension group during the pre-hypertension period, and 3) slopes before and slope after hypertension onset. We set baseline covariates at the most common characteristics: aged 60, female, an education level below NVQ3/GCE A level, married, current drinkers but not smokers, BMI between 24.9 and 29.9, engaging moderate-vigorous activities, non-depressed, and free from chronic disease (diabetes, cancer, chronic lung disease, or heart failure). Random effects were set to be 0.

**Figure 2.**
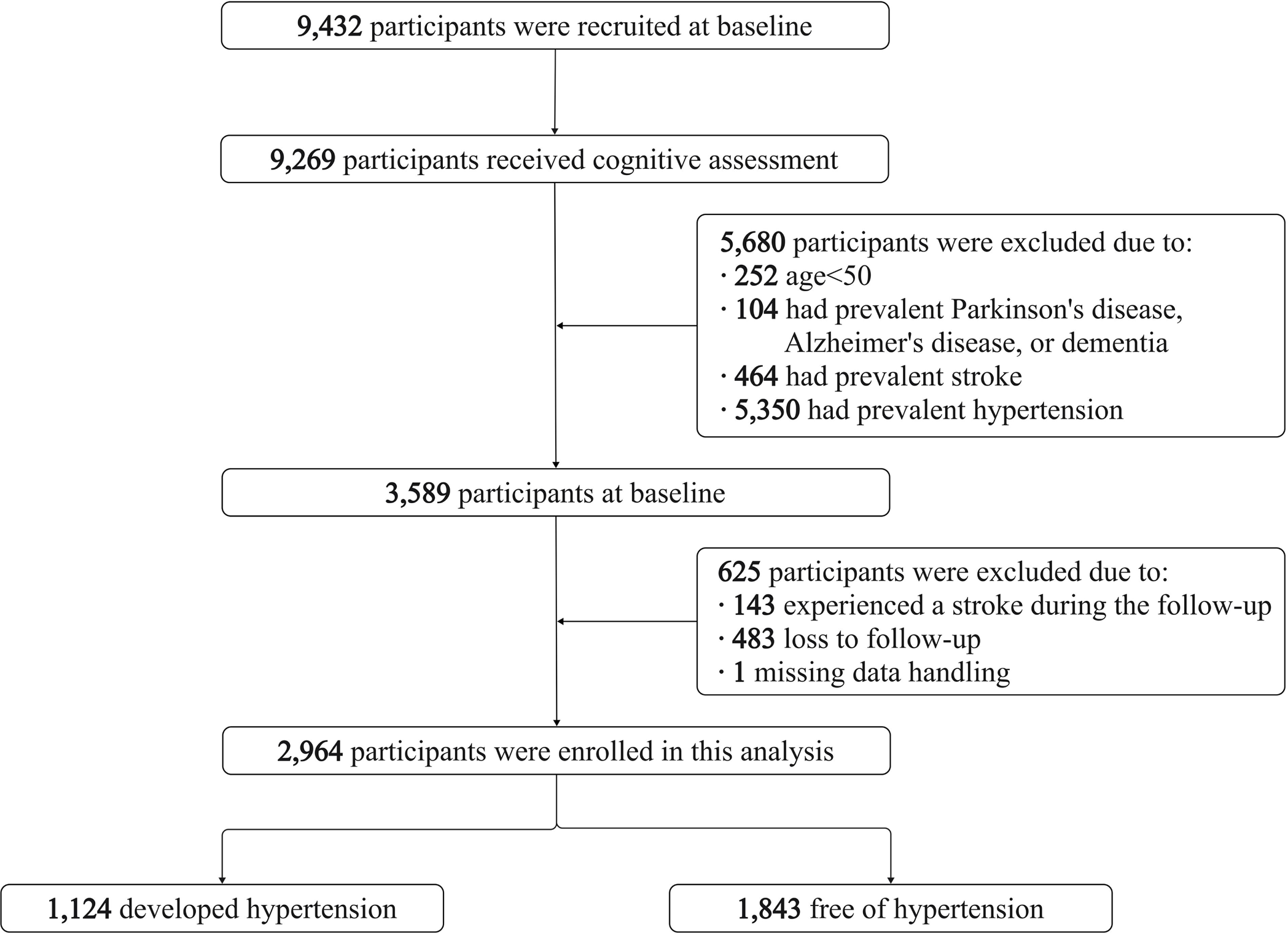
Cognitive z score trajectories calculated by the linear mixed-effects models. The red lines represent the average cognitive trajectories of 1121 participants before and after new-onset hypertension, while the blue lines represent the average cognitive trajectory of 1843 participants who did not develop hypertension. We set the occurrence of hypertension at the end of the fifth year.

In the subgroup analysis, the primary analyses were repeated and stratified by age at hypertension onset, baseline age, blood pressure control, sex, and education level.(23) An additional linear mixed model further compared the difference-in-differences of the cognitive slope between the pre-hypertension period vs. post-hypertension period, comparing older vs. younger. Briefly, it compared whether the accelerated cognitive decline after hypertension differed between these subgroups (Supplemental Figure 2).

We conducted the following sensitivity analysis to test the robustness of our findings: a) applying the IPTW in the linear mixed effects model; b) comparing the mean rate of cognitive decline of the two groups by eliminating ‘post-hypertension cognitive decline’ variable; c) considering ‘acute cognitive change’ at the time of hypertension onset; d) restricting analysis to participants who completed all cognitive tests. Supplemental Method 3 and Supplemental Figure 3 described the reasons and details of the sensitivity analysis.

All *p* values were from 2-sided tests and results were deemed statistically significant at <0.05. Analyses were performed using SAS version 9.4M7 from December 2023 through April 2024.

## Results

### Cognitive assessments and baseline characteristics

The study included 2,964 participants who were free of hypertension at baseline. The median (interquartile range) follow-up time was 13.8 (11.2, 14.1) years for the hypertension group and 12.2 (5.6, 14.0) years for the hypertension-free group. During the follow-up period, 1,121 (37.8%) developed hypertension. Supplemental Table 4 shows the number of new-onset hypertension during each wave. Hypertension occurred at a mean (SD) of 4.6 (3.2) years after baseline. 55.8% of the participants completed cognitive tests in the final wave (Supplemental Table 5). On average, the hypertension group took 4.1 (1.9) cognitive tests before developing hypertension and 2.8 (1.6) tests after. The first post-hypertension interview was conducted 1.2 (0.8) years after hypertension onset.

Compared with those without hypertension, participants who developed hypertension tended to be older, less likely to smoke or have chronic lung disease or cancer, exhibit a higher percentage of obesity and diabetes, and have higher baseline semantic fluency scores, orientation scores, and number of cognitive interviewers during the follow-up (Table 1). After applying IPTW, the two groups were well-matched on baseline characteristics.

**Table 1.**
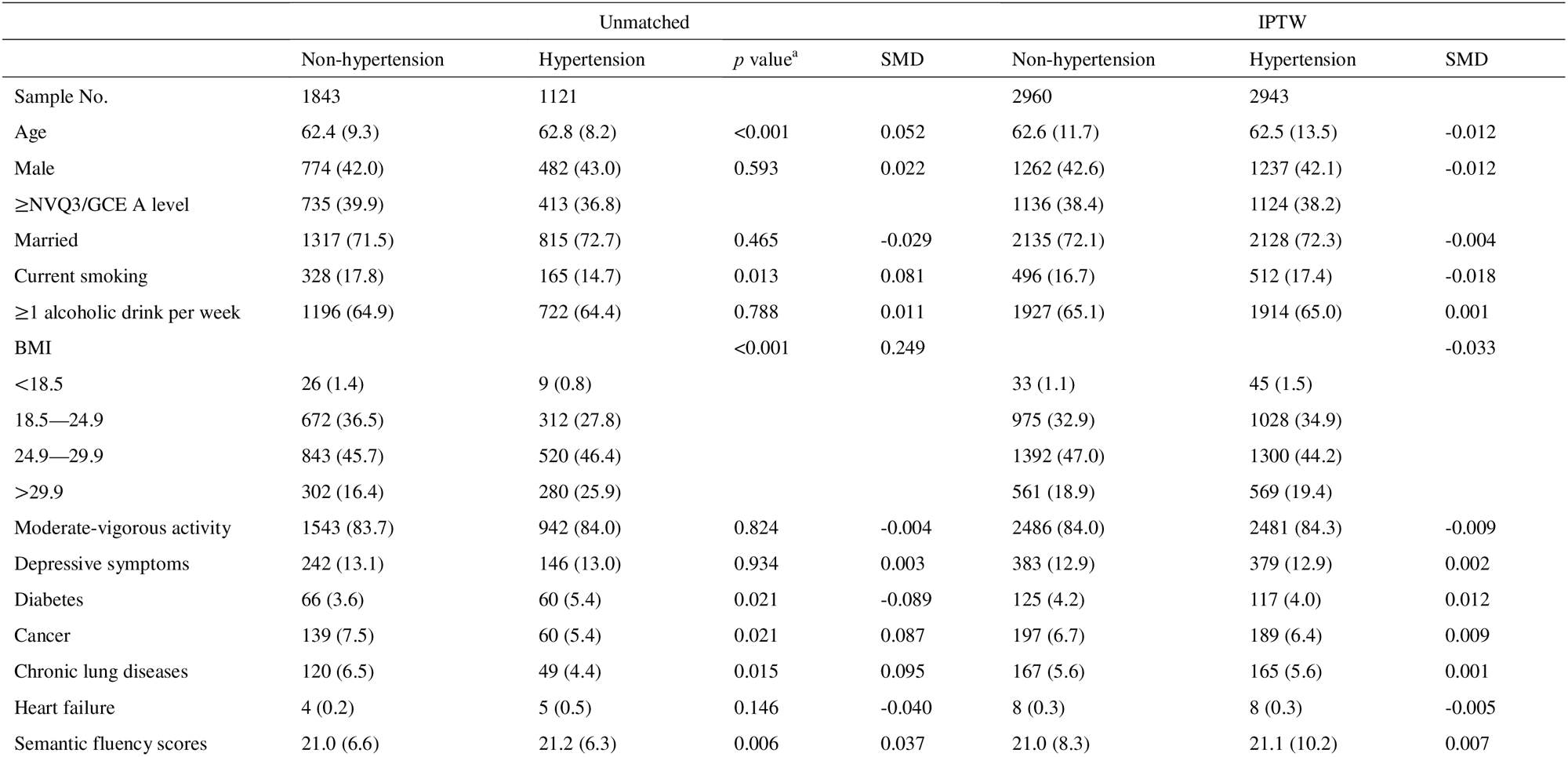
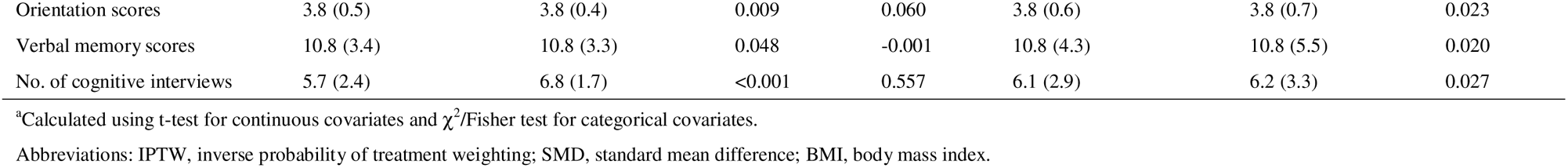
Characteristics according to hypertension onset during the follow-up.

### Primary analysis

For both groups, cognitive scores in all domains gradually declined over time (Figure 2). For the hypertension-free group, global cognition scores declined at a rate of 0.015 SD per year (β, −0.015 SD/year; 95% CI, −0.018 to −0.011; *p*<0.001; Table 2). The hypertension group showed a similar rate of cognitive decline during the pre-hypertension period (difference in global cognition, *p*=0.418). After hypertension onset, the decline rate of global cognition scores was significantly faster than the decline rate before hypertension (β, −0.015 SD/year; 95% CI, −0.026 to −0.003; *p*=0.011), equally twice the rate of normal aging (percentage increase, 100%; 95% CI, 59% to 141%). As shown by Figure 2, 7.5 years after hypertension, the global cognition score of the hypertension group began to be lower than that of the hypertension-free group, despite the hypertension group having a higher score at baseline.

**Table 2.** Results of primary analysis^a^.

| Variables<sup>b</sup> | Global cognition |  | Semantic fluency |  | Orientation |  | Memory |  |
| --- | --- | --- | --- | --- | --- | --- | --- | --- |
| | $\beta$ (95% CI) | *p* | $\beta$ (95% CI) | *p* | $\beta$ (95% CI) | *p* | $\beta$ (95% CI) | *p* |
| Difference in baseline | 0.065 (-0.004, 0.134) | 0.066 | 0.064 (-0.009, 0.136) | 0.084 | 0.056 (-0.022, 0.133) | 0.160 | 0.023 (-0.045, 0.091) | 0.502 |
| Slope of hypertension-free group | -0.015 (-0.018, -0.011) | <0.001 | -0.007 (-0.011, -0.003) | <0.001 | -0.013 (-0.018, -0.008) | <0.001 | -0.023 (-0.027, -0.019) | <0.001 |
| Difference in slope before hypertension | 0.004 (-0.005, 0.012) | 0.418 | 0.003 (-0.006, 0.013) | 0.518 | -0.001 (-0.014, 0.011) | 0.832 | 0.007 (-0.002, 0.016) | 0.117 |
| Changes in slope after hypertension | -0.015 (-0.026, -0.003) | 0.011 | -0.015 (-0.027, -0.003) | 0.017 | -0.012 (-0.028, 0.005) | 0.157 | -0.022 (-0.033, -0.010) | <0.001 |

<sup>a</sup>Adjusted for baseline age, sex, education, marital status, current smoking, current drinking, BMI, physical activity, depression, diabetes, cancer, chronic lung diseases, and heart failure.
<sup>b</sup>Detailed description for variables is shown in Supplemental Figure 1.

Specific cognitive domains showed different patterns of decline. After hypertension onset, scores of the semantic fluency test (β, −0.015 SD/year; 95% CI, −0.027 to −0.003; *p*=0.017; Figure 2) and memory test (β, −0.022 SD/year; 95% CI, −0.033 to −0.010; *p*<0.001) declined more rapidly than the pre-hypertension rate, while the orientation scores did not show a significant change in the rate of decline (β, −0.012 SD/year; 95% CI, −0.028 to 0.005; *p*=0.157). The results remained consistent after applying IPTW (Supplemental Table 6 and Supplemental Figure 4).

### Risk factors for cognitive decline after hypertension

According to age at hypertension onset and sample size, we divided participants with hypertension into three groups. 394 individuals developed hypertension between 55—64 y, 441 between 65—74 y, and 226 between 75—84 y. Compared with participants developing hypertension between 55—64 y, those developing hypertension between 65—74 y experienced more accelerated cognitive decline in global cognition after hypertension (−0.036 SD/year; 95% CI, −0.064 to −0.009; *p*=0.009; Supplemental Table 7 and Figure 3). The oldest age group (75—84 y) showed a faster global cognitive decline before hypertension (−0.032 SD/year; 95% CI, −0.054 to −0.010; *p*=0.005) but did not exhibit a significant change in the rate of decline after hypertension (−0.022 SD/year; 95% CI, −0.055 to 0.010; *p*=0.183). Overall, older age groups were associated with faster cognitive decline in certain cognitive domains. Therefore, younger age at hypertension onset was not associated with a faster cognitive decline either before or after hypertension.

**Figure 3.**
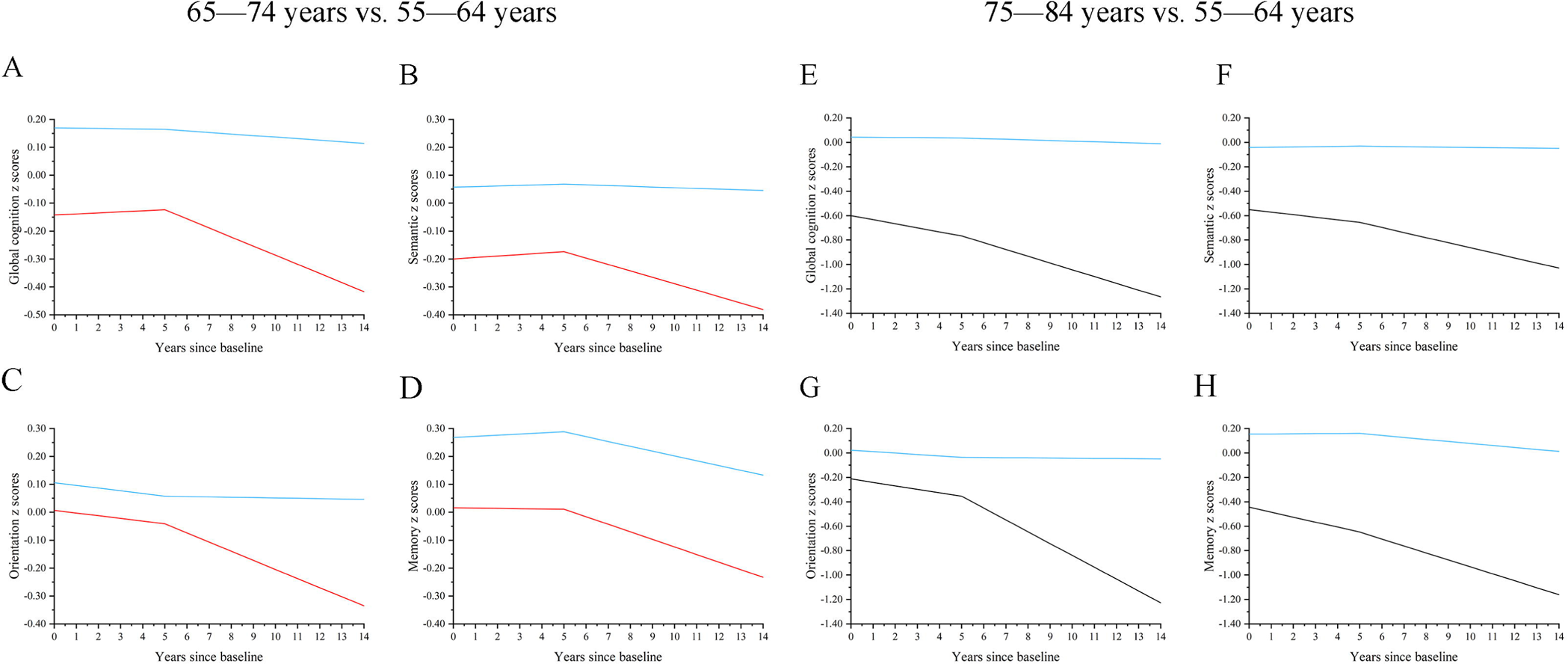
Cognitive z score trajectories according to age at hypertension onset. The blue lines represent the average cognitive trajectories of 394 participants who developed hypertension between ages 55—64 (panels A to H). The red lines represent 441 participants who developed hypertension between ages 65—74 (panels A to D). The black lines represent 226 participants who developed hypertension between ages 75—84 (panels E to H). We set the occurrence of hypertension at the end of the fifth year.

Blood pressure control (≥140/90 vs. 130/80—139/89 mmHg, global cognition, *p* for interaction=0.681; ≥ 140/90 vs. < 130/80 mmHg, global cognition, *p* for interaction=0.989), sex (global cognition, *p* for interaction=0.963), or education level (global cognition, *p* for interaction=0.478) did not significantly modify the effect of hypertension on post-hypertension cognitive decline. Older age was associated with steeper cognitive decline in orientation test (70—99 vs. 50—59, *p* for interaction=0.002), but not in global cognition (*p* for interaction=0.188, Supplemental Tables 8, 9, 10, and 11).

### Sensitivity analysis

While comparing the mean cognitive slope from baseline to the end of the follow-up between the hypertension group and the hypertension-free group, the hypertension group exhibited higher baseline scores in all cognitive domains (global cognition; 0.095 SD; 95% CI, 0.030 to 0.160; *p*=0.004; Supplemental Table 12 and Supplemental Figure 5). Both groups experienced a decline in cognitive scores during the follow-up period. The hypertension group declined faster in domains of memory (β, −0.006 SD/year; 95% CI, −0.012 to −0.000; *p*=0.038). For domains of global cognition (β, −0.006 SD/year; 95% CI, −0.011 to −0.000; *p*=0.050), semantic fluency (β, −0.006 SD/year; 95% CI, −0.012 to −0.000; *p*=0.060), and orientation (β, −0.008 SD/year; 95% CI, −0.016 to 0.001; *p*=0.081), there was no statistically significant difference in mean rates of cognitive decline. The ‘acute cognitive change’ at the time of hypertension onset was not significant (Supplemental Table 13 and Supplemental Figure 6). The primary results were consistent when taking ‘acute cognitive change’ into consideration or restricting participants to those who attended all interviews (Supplemental Tables 13 and 14).

## Discussion

In this longitudinal study, new-onset hypertension was associated with faster cognitive decline in global cognition, semantic fluency, and memory, but not in orientation ability. The rate of cognitive decline among those who later developed hypertension during the pre-hypertension period was similar to the decline rate of those who remained hypertension-free throughout the study. Older participants experienced faster cognitive decline both before and after the onset of hypertension compared to younger participants.

Our findings on global cognition align with existing evidence that hypertension increases the risk of vascular dementia (VaD), AD, and all-cause dementia.(2,13,24) Several studies have investigated the domain-specific cognitive impairments among hypertensive individuals. Executive function enables us to plan, focus attention, and accomplish multiple tasks. In the context of hypertension, the executive function plays a crucial role in the self-management of blood pressure, for example, taking medicine and monitoring blood pressure on time, contacting a doctor for help, treatment of comorbidities, and healthy lifestyles. Although there is a lack of unified evaluation, it is commonly assessed by word fluency test, category fluency test (equivalent to the semantic fluency test in this study), and Trial Making Test A and B.(25) The deterioration of semantic fluency test after hypertension in this study is consistent with previous findings that impaired executive function is the most frequent cognitive change associated with hypertension. Previous studies on memory decline after hypertension have posted mixed results. In a systematic review by Joyce et al., 6 studies reported negative associations between hypertension and memory function, while other 6 studies found no association.(26) Although both negative and non-associations between orientation and hypertension were reported, orientation ability is less frequently studied, perhaps because it less affected by hypertension than other domains.(10,27)

The domain-different results have important implications for understanding the mechanisms of hypertension-related cognitive decline. For instance, researchers who reported non-significant results in the memory domain distinguished changes in brain structure after hypertension from those seen in AD, where memory loss is the earliest symptom. However, the accelerated memory decline observed in our study suggests that AD-related pathologies may play a role in the cognitive deterioration post-hypertension.

The role of age in hypertension-induced cognitive impairment is complex.(28) A small California retirement study reported that participants developing hypertension after 90 years old had a lower risk of dementia than participants developing hypertension between 80—89 y.(29) The Whitehall II cohort study included 8,639 dementia-free participants and discovered that high baseline BP (systolic BP≥130 mmHg) was associated with higher future dementia risk in those with hypertension at age 50, but not at age 60 or 70.(30) The meta-analysis by Ou et al. also concluded that hypertension in midlife was associated with 1.20 to 1.55-fold risk of all-cause dementia and cognitive impairment, while late-life hypertension had a weaker association with dementia (RR, 1.02; 95% CI, 0.94—1.10).(2) However, in the current study, older participants experienced greater post-hypertension cognitive decline. Our findings suggested that older participants might have a higher risk of dementia and their blood pressure should be managed rigorously to prevent dementia. Differences in findings can be ascribed to the following factors. First, dementia is a dichotomous variable, while the cognitive scores were continuous variables. Understanding cognitive decline helps elucidate the mechanisms of dementia onset. Nevertheless, studies analyzing ‘dementia’ and ‘cognitive function’ are not completely equivalent. Second, previous researchers only recruit participants with prevalent hypertension. Studies on incident hypertension are scarce.(31) Third, the definition of hypertension varied. For example, in the previously mentioned Whitehall II study, after including antihypertensive medicine as part of the hypertensive definition, hypertension at age 60 turned out to be significantly associated with dementia.(30) Conversely, some literature supported that older age at hypertension onset was associated with worse cognitive performance or a higher risk of dementia.(32,33) It is worth mentioning that the cognitive declines in older ages are often steeper than those in middle age, following the natural cognitive change pattern.(34)

We excluded participants who experienced a stroke at baseline or during follow-up, indicating that the mechanisms linking hypertension to cognitive decline extend beyond stroke. A primary mechanism is the structural alteration of large, pial and penetrating, and micro cerebral blood vessels, due to factors like atherosclerotic plaques, endothelial dysfunction, inflammation, angiotensin II, and oxidative stress.(35–37) These vascular damages would lead to reduction of cerebral blood flow (CBF), silent stroke, microinfarcts, microbleeds, white matter lesions, and brain atrophy, all of which are important causes of cognitive decline.(38–40) Hypertension itself and reduced CBF also impair metabolite clearance, including amyloid-β (Aβ) clearance, a key player in the pathogenesis of Alzheimer’s disease.(41,42) Additionally, hypertension contributes to VaD and AD through a series of overlapping mechanisms such as impaired regulation ability of CBF, neurovascular coupling, and brain-blood barrier disruption.(43–45) Effective management of hypertension is crucial in mitigating these effects and preserving cognitive health.

To our knowledge, this is the first longitudinal study to track the cognitive change before and after hypertension onset.(31) By using repeated cognitive measurements over a long follow-up period, we illustrated how the hypertension group, despite having better cognitive function at baseline, exhibited worse cognitive function than the hypertension-free group by the end of the follow-up due to post-hypertension cognitive deterioration. The unique study design, where all participants were hypertension-free at baseline, provided new insights into the age difference in cognitive decline following incident hypertension.

Our study, despite this, has several limitations. First, the cognitive test used in ELSA was lack of the ability to diagnose dementia in a valid and consistent manner.(46) We were therefore unable to analyze the changes in dementia probability before and after hypertension.(47) Future studies should use cognitive assessments capable of evaluating both global cognition and diagnosing dementia, such as the Mini-Mental State Examination (MMSE). Second, the study participants of ELSA were primarily White. The generalizability to other population/ethnicity is a concern. Third, some participants were unaware of having hypertension until ELSA measured their blood pressure in Wave 2, 4, 6, or 8, leading to delayed diagnosis. Fourth, cognitive scores did not always decline linearly in cohort studies. Nonetheless, as described in the sensitivity analysis, the hypertension group still showed a tendency towards a faster decline rate after eliminating the ‘post-hypertension cognitive decline’ variable. Our primary results on post-hypertension accelerated decline would not be biased by either non-linearity or diagnosis delay. Fifth, our models captured the population means. The figures only visualized the average cognitive trends of the whole study sample, though individual variations existed. It was inevitable that some individuals’ cognitive function worsened after hypertension while others did not. Sixth, some potential covariates were unavailable even though we have adjusted for a variety of confounders. For instance, the ApoE4 status and the brain imaging data in substudy.

## Conclusion

Our study found that cognitive function declined more rapidly after new-onset hypertension compared with the pre-hypertension period. Participants with younger age at hypertension onset did not show faster cognitive decline. No pre-hypertension cognitive disadvantage was observed. Thus, early antihypertensive treatment in both midlife and late life is an effective strategy to prevent dementia. In the future, well-designed randomized controlled trials are still needed to further understand the effect of hypertension control on brain health.(4,5,48)

## Supporting information

STROBE

## Data Availability

All data produced in the present study are available upon reasonable request to the authors.

## Acknowledgments

The authors thank the U.K. Data Archive for using data from the English Longitudinal Study of Ageing.

## List of abbreviations

AD: Alzheimer’s Disease
BMI: body mass index
BP: blood pressure
CBF: cerebral blood flow
ELSA: English Longitudinal Study of Ageing
IPTW: inverse probability of treatment weight
SMD: standardized mean difference
VaD: vascular dementia.

