## Supplementary figures and images for "Trajectories of Cognitive Decline Before and After New-onset Hypertension"

### Figure 2

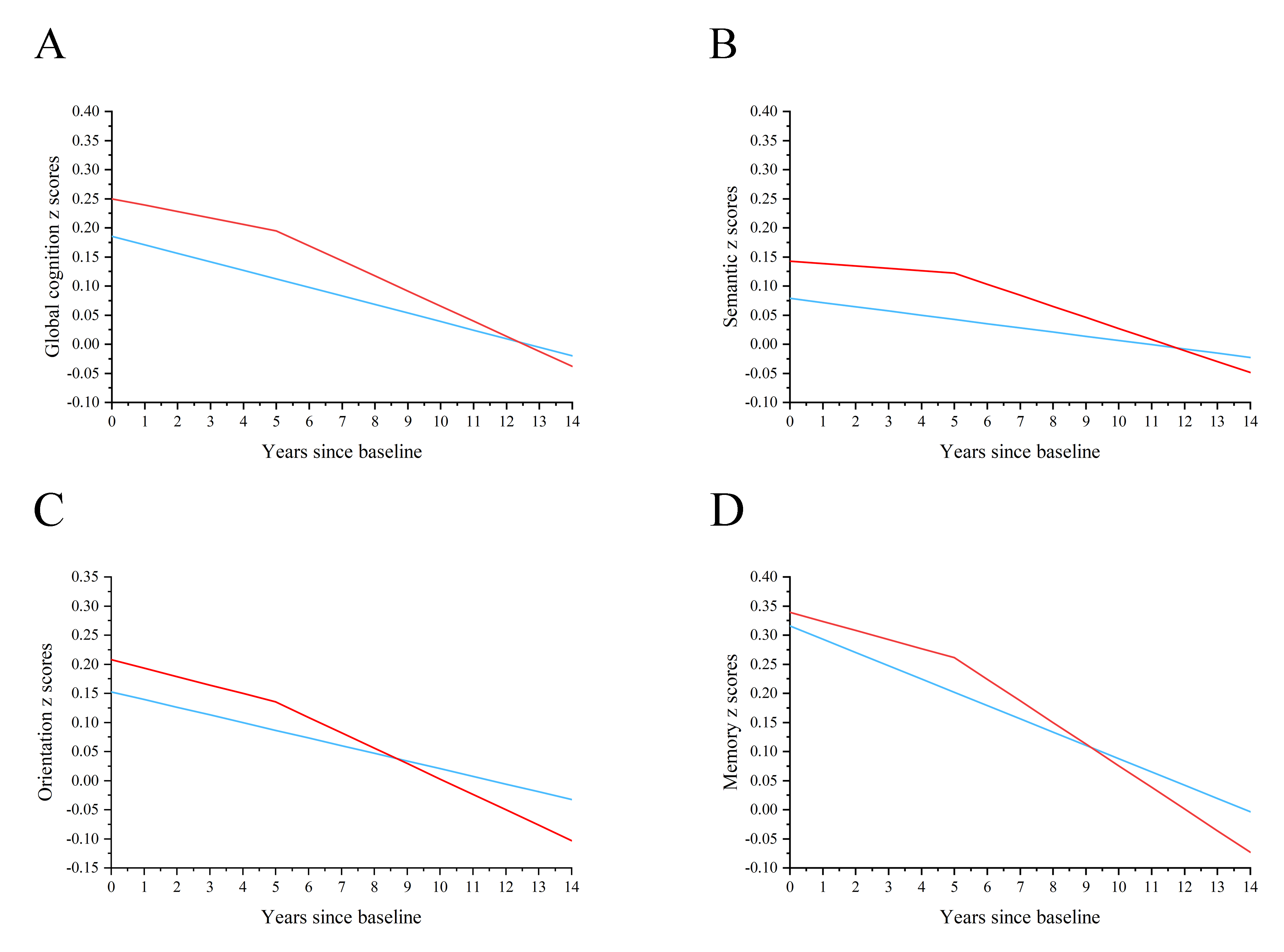
