## Supplemental Appendix for "Trajectories of Cognitive Decline Before and After New-onset Hypertension"

**eAppendix 1**

**Supplemental Table 1.** Comparison of baseline characteristics between participants included and excluded due to loss to follow-up

**Supplemental Method 1.** Covariates

**eMethod2.** Missing data

**Supplemental Table 2.** Results of primary analysis after multiple imputation

**Supplemental Table 3.** Results of primary analysis after excluding participants with missing covariates

**Supplemental Figure 1.** The conceptual model of our main analysis

**Supplemental Figure 2.** The conceptual model of analysis of risk factors

**Supplemental Figure 3.** The conceptual model of analysis of acute cognitive change

**Supplemental Method 3.** Sensitivity analysis

**Supplemental Table 4.** Number of hypertension onset between each wave

**Supplemental Table 5.** Number of available cognition measurements at waves 2 to 9

**Supplemental Table 6.** Results of primary analysis after Inverse Probability of Treatment-Weighting (IPTW)

**Supplemental Figure 4.** Cognitive z score trajectories calculated by the IPTW linear mixed-effects models

**Supplemental Table 7.** Prespecified Subgroup Analysis according to age at hypertension onset

**Supplemental Table 8.** Prespecified Subgroup Analysis according to hypertension control

**Supplemental Table 9.** Subgroup Analysis according to baseline age

**Supplemental Table 10.** Subgroup analysis according to sex

**Supplemental Table 11.** Subgroup analysis according to education

**Supplemental Table 12.** Cognitive z scores trajectories without considering post-hypertension cognitive decline

**Supplemental Figure 5.** Cognitive z score trajectories without considering post-hypertension cognitive decline

**Supplemental Table 13.** Primary analysis after considering ‘acute cognitive change’ at hypertension onset

**Supplemental Figure 6.** Cognitive z score trajectories after considering ‘acute cognitive change’ at hypertension onset

**Supplemental Table 14.** Primary analysis only including participants who attended all cognitive tests (743 in the hypertension-free group and the 623 in hypertension group

**Supplementary Reference**

**Supplemental Table 1. Comparison of baseline characteristics between participants included and excluded due to loss to follow-up**

|  | Included | Loss to follow-up | *p* value^*^ |
| --- | --- | --- | --- |
| Sample No. | 2964 | 481 |  |
| Age | 62.6 (8.9) | 66.7 (11.6) | <0.001 |
| Male | 1256 (42.4) | 211 (43.9) | 0.539 |
| $\geq$NVQ3/GCE A level | 1148 (38.7) | 105 (21.8) | <0.001 |
| Married | 2132 (71.9) | 299 (62.2) | <0.001 |
| Current smoking | 493 (16.7) | 102 (21.7) | 0.008 |
| $\geq$1 alcoholic drink per week | 1918 (64.7) | 241 (50.1) | <0.001 |
| BMI |  |  | <0.001 |
| $<$18.5 | 96 (3.2) | 60 (12.5) |  |
| 18.5—24.9 | 984 (33.2) | 154 (32.0) |  |
| 24.9—29.9 | 1302 (43.9) | 190 (39.5) |  |
| $>$29.9 | 582 (19.6) | 77 (16.0) |  |
| Moderate-vigorous activity | 2485 (83.8) | 336 (69.9) | <0.001 |
| Depressive symptoms | 388 (13.2) | 76 (16.3) | 0.068 |
| Diabetes | 126 (4.3) | 25 (5.2) | 0.347 |
| Cancer | 199 (6.7) | 41 (8.5) | 0.148 |
| Chronic lung diseases | 169 (5.7) | 48 (10.0) | <0.001 |
| Heart failure | 9 (0.3) | 6 (1.3) | 0.010 |
| Semantic fluency scores | 21.0 (6.4) | 18.4 (7.0) | <0.001 |
| Orientation scores | 3.8 (0.5) | 3.6 (0.8) | <0.001 |
| Verbal memory scores | 10.8 (3.3) | 9.1 (3.9) | <0.001 |

^*^Calculated using t-test for continuous covariates and χ^2^/Fisher test for categorical covariates. Continuous variables included age and cognitive test scores.

Abbreviations: IPTW, inverse probability of treatment weighting; SMD, standard mean difference; NVQ, national vocational qualification; GCE, general certificate of education; BMI, body mass index.

**Supplemental Method 1. Covariates**

We defined wave 2 as the baseline of our study. Hence, covariates were derived from wave 2, unless otherwise indicated. Age data were extracted directly from the database. Sex was categorized as male or female. Education level was classified as $\geq$NVQ3/GCE A level or $<$NVQ3/GCE A level. In wave 1, the educational level was recorded under the following classification: no qualification, level 1 national vocational qualification (NVQ) or certificate of secondary education, NVQ2 or general certificate of education (GCE) O-level, NVQ3 or GCE A-level, higher qualification but below degree, degree level or higher or NVQ4/5, or others (e.g. refusal). Wave 2 included additional qualifications obtained since wave 1. Qualifications classified as ≥NVQ3/GCE A level included degree/degree level qualification (including higher degree); teaching qualification; nursing qualifications SRN, SCM, SEN, RGN, RM, RHV, Midwife; HNC/HND, BEC/TEC Higher, BTEC Higher/SCOTECH Higher; City and Guilds Full Technological Certificate; SLC/SCE/SUPE at Higher Grade or Certificate of Sixth Year St; SLC Lower; SUPE Lower or Ordinary; School Certificate or Matric; NVQ Level 4/5; or NVQ Level 3/Advanced level GNVQ. The educational data from waves 1 and 2 were combined. Marital status was categorized as married and not married. Current drinking status was grouped into whether or not to drink 1 alcoholic drink per week.(1) Current smoking status was grouped into yes, no, or missing. Body mass index (BMI) was calculated as weight (kg)/height^2^ (m2). As some participants missed BMI data in wave 2, we used values from wave 0 and wave 4 as the estimates at wave 2. After the above imputation, there were still 120 participants with missing data on BMI. Physical activity was categorized into inactive (no moderate or vigorous activity weekly) and moderate-vigorous activity (at least once a week). Depressive symptoms were measured with the eight-item version of the Center for Epidemiologic Studies Depression Scale (CESD-8), with a score>4 indicating depressive symptoms.(2) Diabetes was defined as self-reported doctor-diagnosed diabetes, use of anti-diabetic medication, or haemoglobin A1c (HbA1c)$\geq$6.5%. Cancer, chronic lung diseases, and heart failure were defined by self-reported physician diagnoses.

**Supplemental Method 2. Missing data**

As depicted in Figure 1, after sample selection, there remained 3589 participants in Wave 2. Among them, 0.8% had missing current smoking status, 1.0% had missing depressive data, and 3.3% had missing BMI data. One participant (0.03%) with missing marital status was excluded (shown by missing data handling in Figure 1). In the primary analysis, smoking status was categorized as yes, no, and unknown (who were the 0.8% with missing smoking data). Missing BMI data and CESD were imputed using the mean values of all participants and then passively converted to their categorical values.

As a sensitivity analysis, we repeated our primary analysis using two additional methods of handling missing data: a) performing multiple imputations for current smoking, depressive symptoms, and BMI. Imputation was performed by SAS. Number of imputed datasets was set to be 10. Smoking status was imputed using logistic regression, while BMI and CESD were imputed using linear regression before converting to categorical values (Supplemental Table 2).(3,4); b) excluding participants with missing data on current smoking, BMI, or depressive symptoms (Supplemental Table 3). Results in Supplemental Tables 2 and 3 were consistent with the primary results shown in Table 2.

**Supplemental Table 2. Results of primary analysis after multiple imputation**^a^

|  | Global cognition |  | Semantic fluency |  | Orientation |  | Memory |  |
| --- | --- | --- | --- | --- | --- | --- | --- | --- |
| Variables^b^ | β (95% CI) | *p* | β (95% CI) | *p* | β (95% CI) | *p* | β (95% CI) | *p* |
| Difference in baseline | 0.063 (-0.007, 0.132) | 0.076 | 0.062 (-0.011, 0.134) | 0.095 | 0.054 (-0.024, 0.131) | 0.174 | 0.021 (-0.047, 0.089) | 0.538 |
| Slope of hypertension-free group | -0.015 (-0.018, -0.011) | <0.001 | -0.007 (-0.011, -0.003) | <0.001 | -0.013 (-0.019, -0.008) | <0.001 | -0.023 (-0.027, -0.019) | <0.001 |
| Difference in slope before hypertension | 0.004 (-0.005, 0.012) | 0.419 | 0.003 (-0.006, 0.013) | 0.519 | -0.001 (-0.014, 0.011) | 0.830 | 0.007 (-0.002, 0.016) | 0.115 |
| Changes in slope after hypertension | -0.015 (-0.026, -0.003) | 0.011 | -0.015 (-0.027, -0.003) | 0.018 | -0.012 (-0.028, 0.005) | 0.159 | -0.022 (-0.033, -0.010) | <0.001 |

^a^Adjusted for baseline age, sex, education, marital status, current smoking, current drinking, BMI, physical activity, depression, diabetes, cancer, chronic lung diseases, and heart failure.

^b^Detailed description for variables is shown in Supplemental Figure 1.

**Supplemental Table 3. Results of primary analysis after excluding participants with missing covariates**^a^

|  | Global cognition |  | Semantic fluency |  | Orientation |  | Memory |  |
| --- | --- | --- | --- | --- | --- | --- | --- | --- |
| Variables^b^ | β (95% CI) | *P* | β (95% CI) | *p* | β (95% CI) | *p* | β (95% CI) | *p* |
| Difference in baseline | 0.054 (-0.016, 0.124) | 0.129 | 0.055 (-0.018, 0.129) | 0.139 | 0.048 (-0.030, 0.125) | 0.232 | 0.021 (-0.047, 0.090) | 0.540 |
| Slope of hypertension-free group | -0.015 (-0.019, -0.011) | <0.001 | -0.008 (-0.011, -0.004) | <0.001 | -0.014 (-0.019, -0.008) | <0.001 | -0.023 (-0.026, -0.019) | <0.001 |
| Difference in slope before hypertension | 0.004 (-0.005, 0.013) | 0.339 | 0.004 (-0.006, 0.013) | 0.443 | -0.001 (-0.014, 0.012) | 0.889 | 0.007 (-0.002, 0.016) | 0.142 |
| Changes in slope after hypertension | -0.015(-0.027, -0.003) | 0.011 | -0.014 (-0.026, -0.002) | 0.026 | -0.013 (-0.030, 0.004) | 0.127 | -0.021 (-0.033, -0.010) | <0.001 |

^a^Adjusted for baseline age, sex, education, marital status, current smoking, current drinking, BMI, physical activity, depression, diabetes, cancer, chronic lung diseases, and heart failure.

^b^Detailed description for variables is shown in Supplemental Figure 1.

**Supplemental Figure 1.** **The conceptual model of our main analysis.**

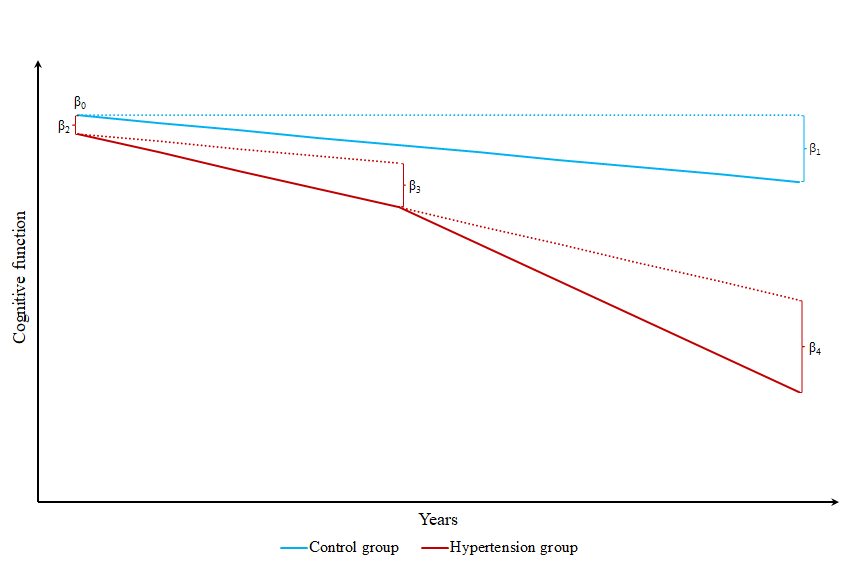

The conceptual model of our study. Time on the x-axis is the years from the date of the first cognitive test. The Y-axis represents the cognitive function. The blue line represents the possible trajectory of the control group without hypertension. We hypothesized their cognitive function declined annually. The cognitive trajectories of the hypertension group (red lines) consisted of the trajectories before hypertension and the accelerated decline after hypertension.

$\beta$0: The predicted cognition of the without-hypertension group at time t=0.

$\beta$1: The difference in cognition from time t to time t+1 in the without-hypertension group, representing the average cognitive slope of the entire without-hypertension group.

$\beta$2: The difference in cognition at time t=0 in the hypertension group compared to the without-hypertension group. Its effect size was named ‘difference in baseline’ in Table 2.

$\beta$3: The difference in slope in the hypertension group compared to the without-hypertension group in the pre-hypertension period.

$\beta$4: The change in slope in the post-hypertension period compared to the pre-hypertension period. After hypertension, we hypothesized the cognitive decline rate was combined with the pre-hypertension decline rate and an accelerated decline caused by hypertension. We assumed hypertension affects cognition in all years after hypertension.

**Supplemental Figure 2. The conceptual model of analysis of risk factors.**

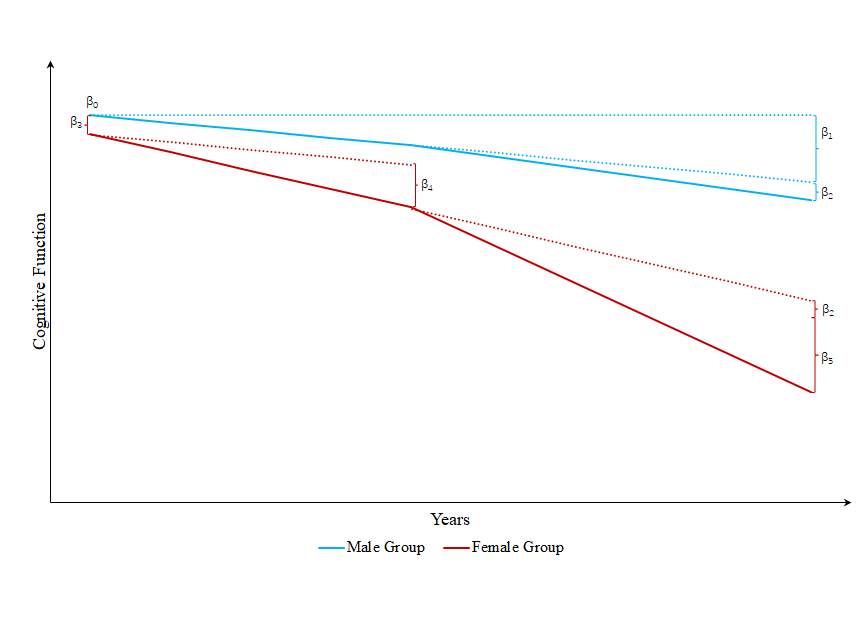

This model examines the effect of risk factors (e.g. female sex) on the effect of hypertension on cognitive trajectories. Time on the x-axis is the years from the date of the first cognitive test. The Y-axis is the cognitive function. The blue line represents the possible trajectory of the male participants (reference group) with hypertension onset. While the red line represents the possible trajectory of the female participants with hypertension onset.

$\beta$0: The predicted cognition of the male group at time t=0.

$\beta$1: The difference in cognition from time t to time t+1 among the male group.

$\beta$2: The change in slope in the post-hypertension period compared to the pre-hypertension period among the male group.

$\beta$3: The difference in cognition at time t=0 in the female group compared to the male group.

$\beta$4: The difference in slope in the female group compared to the male group in the pre-hypertension period.

$\beta$5: The effect of female sex on the effect of hypertension on the changes in cognitive slope after hypertension onset. The change in slope in the post-hypertension period compared to the pre-hypertension period among the female group equals $\beta$2$+\beta$5. The *p* value for $\beta$5 was named ‘*p* for interaction’ in this study (e.g. Supplemental Tables 7, 8, 9, 10, 11).

**Supplemental Figure 3. The conceptual model of analysis of acute cognitive change.**

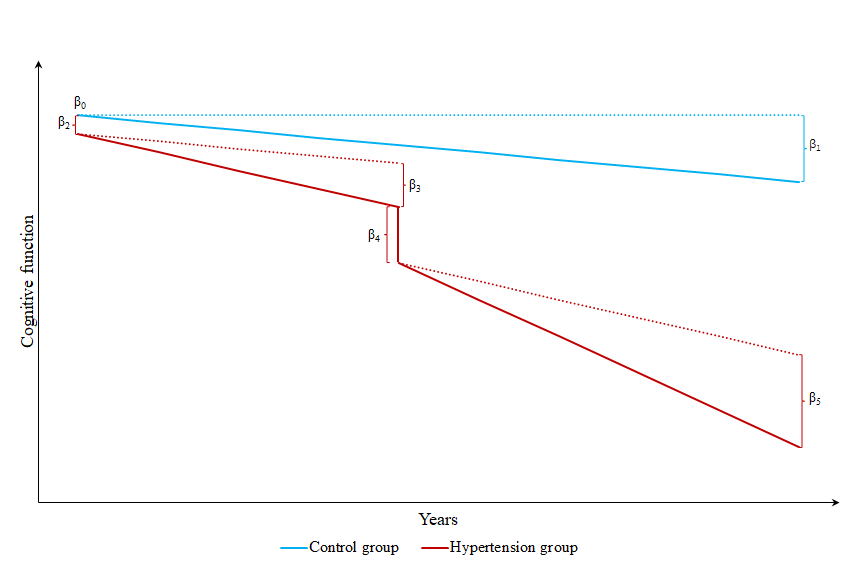

Supplemental Figure 3 is based on Supplemental Figure 1 and adds an ‘acute cognitive change represented by $\beta$4.

$\beta$4: The ‘acute cognitive change’ at the time of hypertension among the hypertension group, was measured by the first post-hypertension cognitive score minus the last pre-hypertension cognitive score. The ‘hypertension status’ was a time-varying dichotomous variable that changed from ‘0’ to ‘1’ at the time of hypertension onset and reflected the acute cognitive change at the time of hypertension. In linear mixed models, we added a fixed effect for ‘hypertension status’ to estimate the effect of ‘acute cognitive change’.

**Supplemental Method 3. Sensitivity analysis**

a) We estimated propensity scores (absolute probability of developing hypertension during the follow-up) using logistic regression. The logistic regression model incorporated all the covariates listed in Table 1, which consisted of covariates, baseline cognitive scores, and No. of cognitive interviews. We included No. of interviews during the follow-up because the hypertension group had a longer follow-up time. We used propensity scores to generate for each participant an inverse probability of treatment weight (IPTW) and then repeated the primary analysis using the IPTW linear mixed model (Supplemental Table 6), which upweighted participants who developed hypertension but appeared to be appropriate control group (without hypertension) candidates and the converse. The IPTW therefore helps balance the characteristics and simulate a randomized trial.

b) Our manuscript compared the control groups’ cognitive slope, the pre-hypertension slope, and the post-hypertension slope, reporting that participants experienced faster cognitive decline in a few years after new-onset hypertension. The ‘post-hypertension cognitive decline’ might be challenged by non-linearity, meaning that cognitive decline from years 7—14 was faster than the decline from years 0—7. Anytime compare a participant’s cognitive trend in the latter years with that in the early years, studies might achieve constant conclusions. To address potential non-linearity, we compared the mean rate of cognitive decline from baseline to follow-up end between the two groups. We used a linear mixed model, fitting fixed effects for intercept, time (years), hypertension (yes or no), hypertension$\times$time interaction, and all covariates, and fitting random effects for intercept and time. If the effect size of ‘hypertension$\times$time interaction’ was significantly (*p*$<$0.05) negative, we concluded that the hypertension group had a faster cognitive decline. Even if the value was between 0.05 and 0.2, there was a tendency for the hypertension group to show a faster decline. If the *p-value* was larger or the effect size of the ‘hypertension$\times$time interaction’ was positive, then the hypertension group did not decline faster than the control group. In this case, the ‘post-hypertension accelerated cognitive decline’ was a false positive.

c) Previous studies reported that stroke patients experienced an ‘acute cognitive decline’ at the time of stroke, after considering the cognition of the without-stroke group, the cognitive trends before stroke, and the cognitive trends after stroke. The ‘acute cognitive decline’ aligns with the phenomenon that numerous neurologists reported that the risk of dementia onset is quite high within one year after stroke.(5,6) Therefore, we examined whether our hypertension participants experienced an ‘acute cognitive decline’ through methods illustrated in Supplemental Figure 3.

d) A method to handle missing data. An unavoidable selection bias of the cognitive study is ‘about to receive cognitive test’. Those without cognitive test data must be unhealthier or even dead (Supplemental Table 1). These kinds of studies always focus on relatively healthy participants, unless the interviewers recorded narratively why a participant did not finish a cognitive test. One solution is comparing the main results with the results of participants who receive all cognitive tests (healthier). This analysis also addresses the problem of different follow-up times between the two groups.

**Supplemental Table 4. Number of hypertension onset between each wave**

| Time of hypertension onset | No. |
| --- | --- |
| Wave 2 to Wave 3 | 179 |
| Wave 3 to Wave 4 | 375 |
| Wave 4 to Wave 5 | 205 |
| Wave 5 to Wave 6 | 97 |
| Wave 6 to Wave 7 | 89 |
| Wave 7 to Wave 8 | 121 |
| Wave 8 to Wave 9 | 55 |

**Supplemental Table 5. Number of available cognition measurements at waves 2 to 9**

| **Waves** | **Non-hypertension (%)** | **Hypertension (%)** | **Total (%)** |
| --- | --- | --- | --- |
| Wave 2 (2004—2005) | 1843 (100.0)^*^ | 1121 (100.00) | 2964 (100.0) |
| Wave 3 (2006—2007) | 1742 (95.5) | 1071 (95.5) | 2813 (94.9) |
| Wave 4 (2008—2009) | 1458 (79.1) | 1058 (94.4) | 2516 (84.9) |
| Wave 5 (2010—2011) | 1357 (73.6) | 1040 (92.8) | 2397 (80.9) |
| Wave 6 (2012—2013) | 1248 (67.7) | 1007 (89.8) | 2255 (76.1) |
| Wave 7 (2014—2015) | 1138 (61.8) | 912 (81.4) | 2050 (69.1) |
| Wave 8 (2016—2017) | 994 (53.9) | 824 (73.5) | 1818 (61.3) |
| Wave 9 (2018—2019) | 905 (49.1) | 749 (66.8) | 1654 (55.8) |

^*^No. (percent of Wave 2).

**Supplemental Table 6. Results of primary analysis after Inverse Probability of Treatment-Weighting (IPTW)**^a^

|  | Global cognition |  | Semantic fluency |  | Orientation |  | Memory |  |
| --- | --- | --- | --- | --- | --- | --- | --- | --- |
| Variables^b^ | β (95% CI) | *p* | β (95% CI) | *p* | β (95% CI) | *p* | β (95% CI) | *p* |
| Difference in baseline | 0.046 (-0.022, 0.114) | 0.182 | 0.043 (-0.028, 0.114) | 0.237 | 0.024 (-0.050, 0.098) | 0.524 | 0.012 (-0.055, 0.079) | 0.725 |
| Slope of hypertension-free group | -0.015 (-0.019, -0.011) | <0.001 | -0.008 (-0.012, -0.004) | <0.001 | -0.014 (-0.020, -0.009) | <0.001 | -0.023 (-0.027, -0.019) | <0.001 |
| Difference in slope before hypertension | 0.004 (-0.005, 0.012) | 0.421 | 0.004 (-0.005, 0.013) | 0.394 | -0.001 (-0.013, 0.011) | 0.845 | 0.006 (-0.003, 0.015) | 0.187 |
| Changes in slope after hypertension | -0.015 (-0.026, -0.003) | 0.013 | -0.015 (-0.028, -0.003) | 0.014 | -0.011 (-0.028, 0.005) | 0.181 | -0.021 (-0.033, -0.010) | <0.001 |

^a^Adjusted for baseline age, sex, education, marital status, current smoking, current drinking, BMI, physical activity, depression, diabetes, cancer, chronic lung diseases, and heart failure.

^b^Detailed description for variables is shown in Supplemental Figure 1.

**Supplemental Figure 4. Cognitive z score trajectories calculated by the IPTW linear mixed-effects models.** The figure was fitted based on the analysis in Supplemental Table 6. The red lines represent the average cognitive trajectories before and after new-onset hypertension, while the blue lines represent the average cognitive trajectory of participants who did not develop hypertension. We set the occurrence of hypertension at the end of the fifth year.

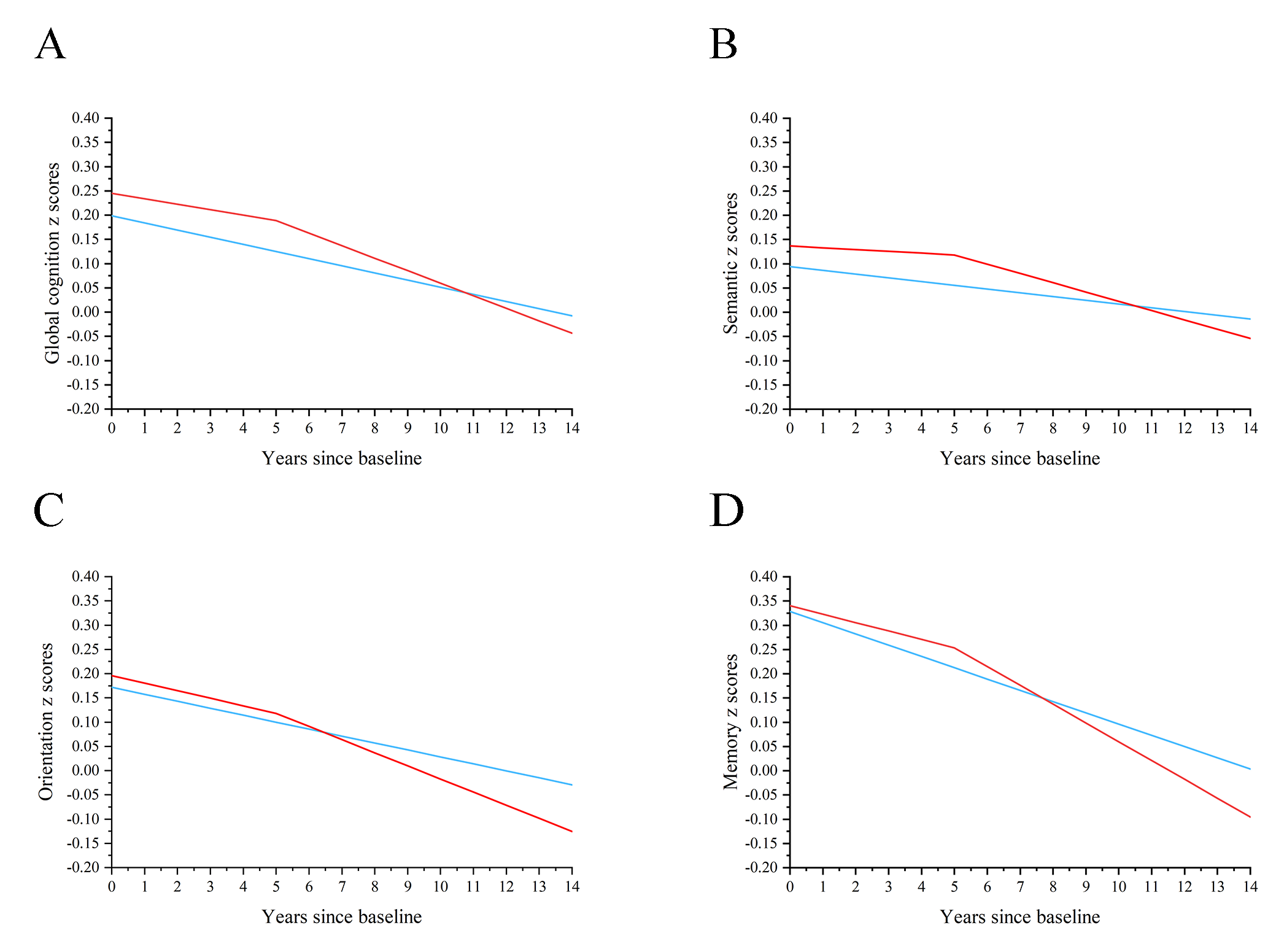

**Supplemental Table 7. Prespecified Subgroup Analysis according to age at hypertension onset**^a^

|  | Global cognition |  | Semantic fluency |  | Orientation |  | Memory |  |
| --- | --- | --- | --- | --- | --- | --- | --- | --- |
| Variables^b^ | β (95% CI) | *p* | β (95% CI) | *p* | β (95% CI) | *p* | β (95% CI) | *p* |
| Group 1, reference (55—64, n$=$394) |  |  |  |  |  |  |  |  |
| Slope before hypertension | -0.001 (-0.018, 0.016) | 0.907 | 0.002 (-0.016, 0.021) | 0.820 | -0.010 (-0.032, 0.013) | 0.396 | 0.004 (-0.013, 0.021) | 0.639 |
| Changes in slope after hypertension | -0.005 (-0.026, 0.017) | 0.676 | -0.005 (-0.028, 0.019) | 0.695 | 0.008 (-0.019, 0.036) | 0.543 | -0.021 (-0.042, 0.000) | 0.048 |
| Group 2 (65—74, n$=$441) vs. Group 1 |  |  |  |  |  |  |  |  |
| Difference in pre-hypertension slope | 0.005 (-0.016, 0.025) | 0.644 | 0.003 (-0.019, 0.025) | 0.782 | 0.000 (-0.027, 0.027) | 0.999 | -0.005 (-0.025, 0.015) | 0.635 |
| Difference in changes in slope after hypertension | -0.036 (-0.064, -0.009) | 0.009 | -0.028 (-0.058, 0.001) | 0.063 | -0.023 (-0.057, 0.012) | 0.192 | -0.026 (-0.053, 0.000) | 0.053 |
| Group 3 (75—84, n$=$226) vs. Group 1 |  |  |  |  |  |  |  |  |
| Difference in pre-hypertension slope | -0.032 (-0.054, -0.010) | 0.005 | -0.023 (-0.046, 0.000) | 0.052 | -0.017 (-0.053, 0.020) | 0.374 | -0.042 (-0.064, -0.019) | <0.001 |
| Difference in changes in slope after hypertension | -0.022 (-0.055, 0.010) | 0.183 | -0.020 (-0.055, 0.014) | 0.246 | -0.069 (-0.119, -0.019) | 0.007 | -0.016 (-0.049, 0.016) | 0.322 |

^a^Adjusted for baseline age, sex, education, marital status, current smoking, current drinking, BMI, physical activity, depression, diabetes, cancer, chronic lung diseases, and heart failure.

^b^Detailed description for variables is shown in Supplemental Figure 2.

**Supplemental Table 8. Prespecified Subgroup Analysis according to hypertension control**^a^

|  | Global cognition |  | Semantic fluency |  | Orientation |  | Memory |  |
| --- | --- | --- | --- | --- | --- | --- | --- | --- |
| Variables^b^ | β (95% CI) | *p* | β (95% CI) | *p* | β (95% CI) | *p* | β (95% CI) | *p* |
| Group 1, reference ($\geq$140/90 mmHg, n$=$493) |  |  |  |  |  |  |  |  |
| Slope before hypertension | -0.007 (-0.021, 0.008) | 0.370 | -0.001 (-0.015, 0.013) | 0.895 | -0.025 (-0.049, -0.001) | 0.042 | -0.009 (-0.024, 0.006) | 0.243 |
| Changes in slope after hypertension | -0.015 (-0.034, 0.004) | 0.116 | -0.011 (-0.031, 0.008) | 0.256 | -0.003 (-0.035, 0.029) | 0.855 | -0.028 (-0.047, -0.009) | 0.005 |
| Group 2 (130/80—139/89 mmHg, n$=$314) vs. Group 1 | | | | | | | | |
| Difference in pre-hypertension slope | -0.005 (-0.024, 0.014) | 0.603 | -0.002 (-0.022, 0.018) | 0.828 | 0.013 (-0.017, 0.044) | 0.389 | -0.006 (-0.025, 0.012) | 0.516 |
| Difference in changes in slope after hypertension | -0.006 (-0.034, 0.022) | 0.681 | -0.011 (-0.041, 0.019) | 0.461 | -0.007 (-0.050, 0.035) | 0.734 | -0.004 (-0.031, 0.023) | 0.773 |
| Group 3 ($<$130/80 mmHg, n$=$196) vs. Group 1 |  |  |  |  |  |  |  |  |
| Difference in pre-hypertension slope | -0.005 (-0.027, 0.017) | 0.646 | -0.006 (-0.028, 0.016) | 0.590 | 0.018 (-0.019, 0.055) | 0.352 | -0.009 (-0.032, 0.013) | 0.428 |
| Difference in changes in slope after hypertension | 0.000 (-0.032, 0.031) | 0.989 | 0.002 (-0.030, 0.033) | 0.909 | -0.048 (-0.101, 0.005) | 0.078 | 0.018 (-0.013, 0.049) | 0.266 |

^a^Adjusted for baseline age, sex, education, marital status, current smoking, current drinking, BMI, physical activity, depression, diabetes, cancer, chronic lung diseases, and heart failure.

^b^Detailed description for variables is shown in Supplemental Figure 2.

**Supplemental Table 9. Subgroup Analysis according to baseline age^a^**

|  | Group 1 (50—59, Reference)^b^ | Group 2 (60—69) | *p* for interaction^c^ | Group 3 (70—99) | *p* for interaction |
| --- | --- | --- | --- | --- | --- |
| Global cognition | -0.009 (-0.024, 0.005) | -0.011 (-0.029, 0.008) | 0.942 | -0.032 (-0.061,-0.003) | 0.188 |
| Semantic fluency | -0.011 (-0.028, 0.005) | -0.005 (-0.025, 0.015) | 0.577 | -0.038 (-0.067, -0.009) | 0.082 |
| Orientation | 0.002 (-0.014, 0.017) | -0.005 (-0.032, 0.022) | 0.893 | -0.074 (-0.136, -0.013) | 0.002 |
| Memory | -0.021 (-0.036, -0.006) | -0.023 (-0.042, -0.005) | 0.937 | -0.031 (-0.060, -0.001) | 0.815 |

^a^Adjusted for baseline age, education, marital status, current smoking, current drinking, BMI, physical activity, depression, diabetes, cancer, chronic lung diseases, and heart failure.

^b^Results represented the accelerated cognitive decline after hypertension. Calculated by the linear mixed model of the primary analysis.

^c^Results of each subgroup were calculated by the model in Supplemental Figure 1, while *p* for interaction was calculated by the model in Supplemental Figure 2.

**Supplemental Table 10. Subgroup analysis according to sex^a^**

|  | Male^b^ | Female | *p* for interaction^c^ |
| --- | --- | --- | --- |
| Global cognition | -0.015 (-0.033, 0.003) | -0.015 (-0.029, 0.000) | 0.963 |
| Semantic fluency | -0.010 (-0.029, 0.009) | -0.019 (-0.035, -0.003) | 0.440 |
| Orientation | 0.018 (-0.005, 0.041) | -0.025 (-0.048, -0.002) | 0.063 |
| Memory | -0.027 (-0.044, -0.010) | -0.018 (-0.033, -0.003) | 0.328 |

^a^Adjusted for baseline age, education, marital status, current smoking, current drinking, BMI, physical activity, depression, diabetes, cancer, chronic lung diseases, and heart failure.

^b^Results represented the accelerated cognitive decline after hypertension. Calculated by the linear mixed model of the primary analysis.

^c^Results of each subgroup were calculated by the model in Supplemental Figure 1, while *p* for interaction was calculated by the model in Supplemental Figure 2.

**Supplemental Table 11. Subgroup analysis according to education^a^**

|  | $<$ NVQ3/GCE A level^b^ | $\geq$NVQ3/GCE A level | *p* for interaction^c^ |
| --- | --- | --- | --- |
| Global cognition | -0.011 (-0.026, 0.003) | -0.021 (-0.039, -0.002) | 0.478 |
| Semantic fluency | -0.015 (-0.030, 0.001) | -0.015 (-0.005, 0.035) | 0.925 |
| Orientation | -0.014 (-0.037, 0.008) | -0.008 (-0.033, 0.017) | 0.567 |
| Memory | -0.017 (-0.032, -0.003) | -0.027 (-0.046, -0.009) | 0.367 |

^a^Adjusted for baseline age, sex, marital status, current smoking, current drinking, BMI, physical activity, depression, diabetes, cancer, chronic lung diseases, and heart failure.

^b^Results represented the accelerated cognitive decline after hypertension. Calculated by the linear mixed model of the primary analysis.

^c^Results of each subgroup were calculated by the model in Supplemental Figure 1, while *p* for interaction was calculated by the model in Supplemental Figure 2.

**Supplemental Table 12. Cognitive z scores trajectories without considering post-hypertension cognitive decline^a^**

|  | Global cognition |  | Semantic fluency |  | Orientation |  | Memory |  |
| --- | --- | --- | --- | --- | --- | --- | --- | --- |
| Variables^b^ | β (95% CI) | *p* | β (95% CI) | *p* | β (95% CI) | *p* | β (95% CI) | *p* |
| Difference in baseline | 0.095 (0.030, 0.160) | 0.004 | 0.093 (0.025, 0.161) | 0.008 | 0.073 (0.002, 0.145) | 0.044 | 0.069 (0.006, 0.132) | 0.033 |
| Slope of hypertension-free group | -0.015 (-0.018, -0.011) | <0.001 | -0.007 (-0.011, -0.003) | <0.001 | -0.013 (-0.019, -0.008) | <0.001 | -0.023 (-0.027, -0.019) | <0.001 |
| Difference in slope | -0.006 (-0.011, 0.000) | 0.050 | -0.006 (-0.012, 0.000) | 0.060 | -0.008 (-0.016, 0.001) | 0.081 | -0.006 (-0.012, 0.000) | 0.038 |

^a^Adjusted for baseline age, sex, education, marital status, current smoking, current drinking, BMI, physical activity, depression, diabetes, cancer, chronic lung diseases, and heart failure.

**Supplemental Figure 5. Cognitive z score trajectories without considering post-hypertension cognitive decline.** The figure was fitted based on the analysis in Supplemental Table 12. The red lines represent the average cognitive trajectory from baseline to follow-up end of 1121 participants who developed hypertension, while the blue lines represent the average cognitive trajectory of 1843 participants without hypertension.

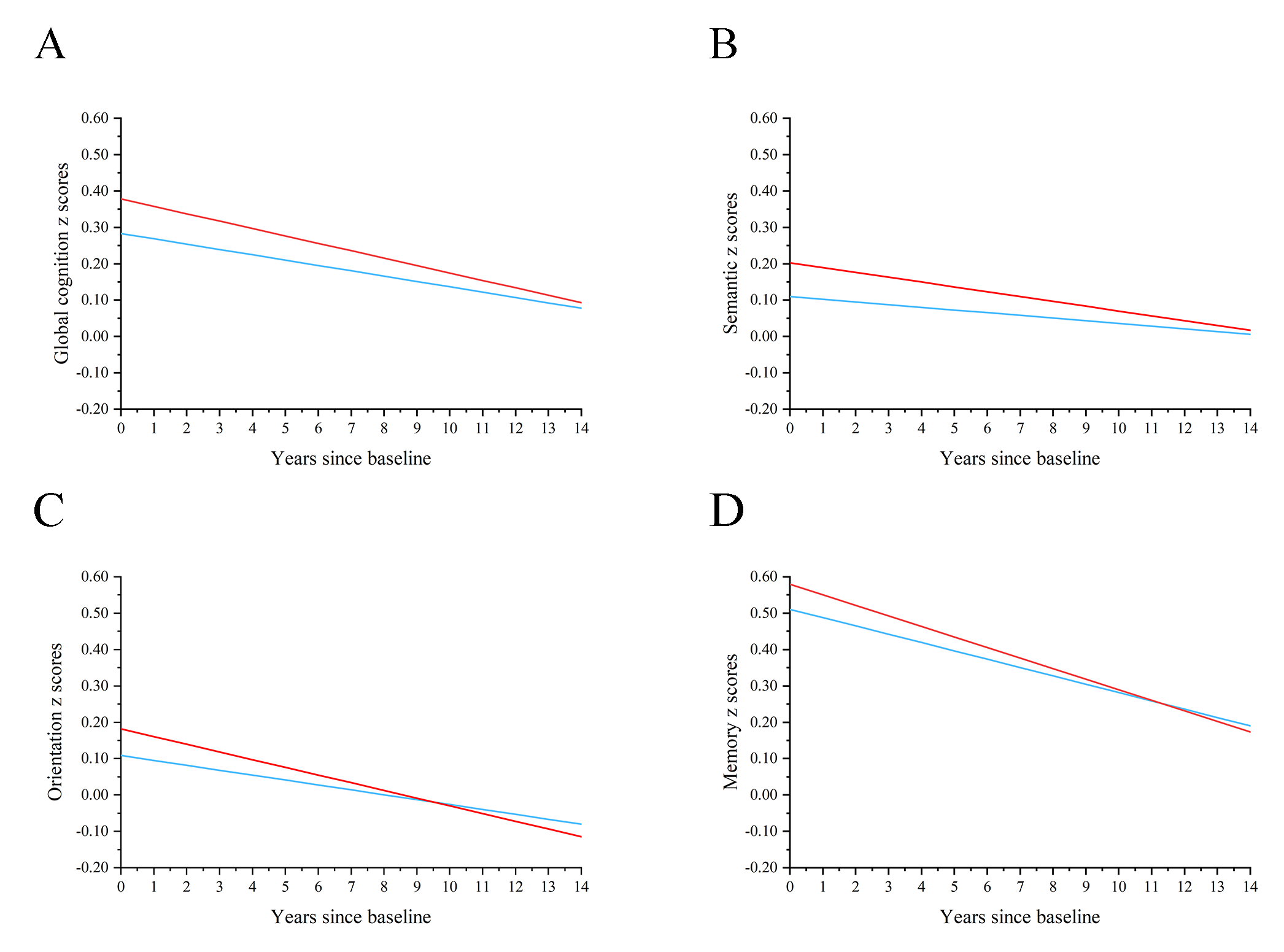

**Supplemental Table 13. Primary analysis after considering ‘acute cognitive change’ at hypertension onset^a^**

|  | Global cognition |  | Semantic fluency |  | Orientation |  | Memory |  |
| --- | --- | --- | --- | --- | --- | --- | --- | --- |
| Variables^b^ | β (95% CI) | *p* | β (95% CI) | *p* | β (95% CI) | *p* | β (95% CI) | *p* |
| Difference in baseline | 0.064 (-0.005, 0.133) | 0.068 | 0.064 (-0.009, 0.136) | 0.086 | 0.055 (-0.022, 0.133) | 0.161 | 0.024 (-0.044, 0.091) | 0.497 |
| Slope of hypertension-free group | -0.015 (-0.018, -0.011) | <0.001 | -0.007 (-0.011, -0.003) | <0.001 | -0.013 (-0.019, -0.008) | <0.001 | -0.023 (-0.027, -0.019) | <0.001 |
| Difference in slope before hypertension | 0.002 (-0.008, 0.012) | 0.678 | 0.001 (-0.009, 0.012) | 0.786 | -0.003 (-0.017, 0.010) | 0.623 | 0.005 (-0.005, 0.015) | 0.334 |
| Changes in slope after hypertension | -0.015 (-0.026, -0.003) | 0.013 | -0.015 (-0.027, -0.002) | 0.019 | -0.013 (-0.029, 0.004) | 0.140 | -0.021 (-0.033, -0.010) | <0.001 |
| Acute cognitive change | 0.017 (-0.032, 0.066) | 0.499 | 0.019 (-0.035, 0.074) | 0.489 | 0.029 (-0.042, 0.099) | 0.423 | 0.027 (-0.023, 0.077) | 0.295 |

^a^Adjusted for baseline age, sex, education, marital status, current smoking, current drinking, BMI, physical activity, depression, diabetes, cancer, chronic lung diseases, and heart failure.

^b^Detailed description of ‘acute cognitive change’ is shown in Supplemental Figure 3.

**Supplemental Figure 6. Cognitive z score trajectories after considering ‘acute cognitive change’ at hypertension onset.** The figure was fitted based on the analysis in Supplemental Table 13. The red lines represent the average cognitive trajectories of 1121 participants before hypertension, an acute cognitive change at the time of hypertension onset, and after hypertension, while the blue lines represent the average cognitive trajectory of 1843 participants who did not develop hypertension. We set the occurrence of hypertension at the end of the fifth year.

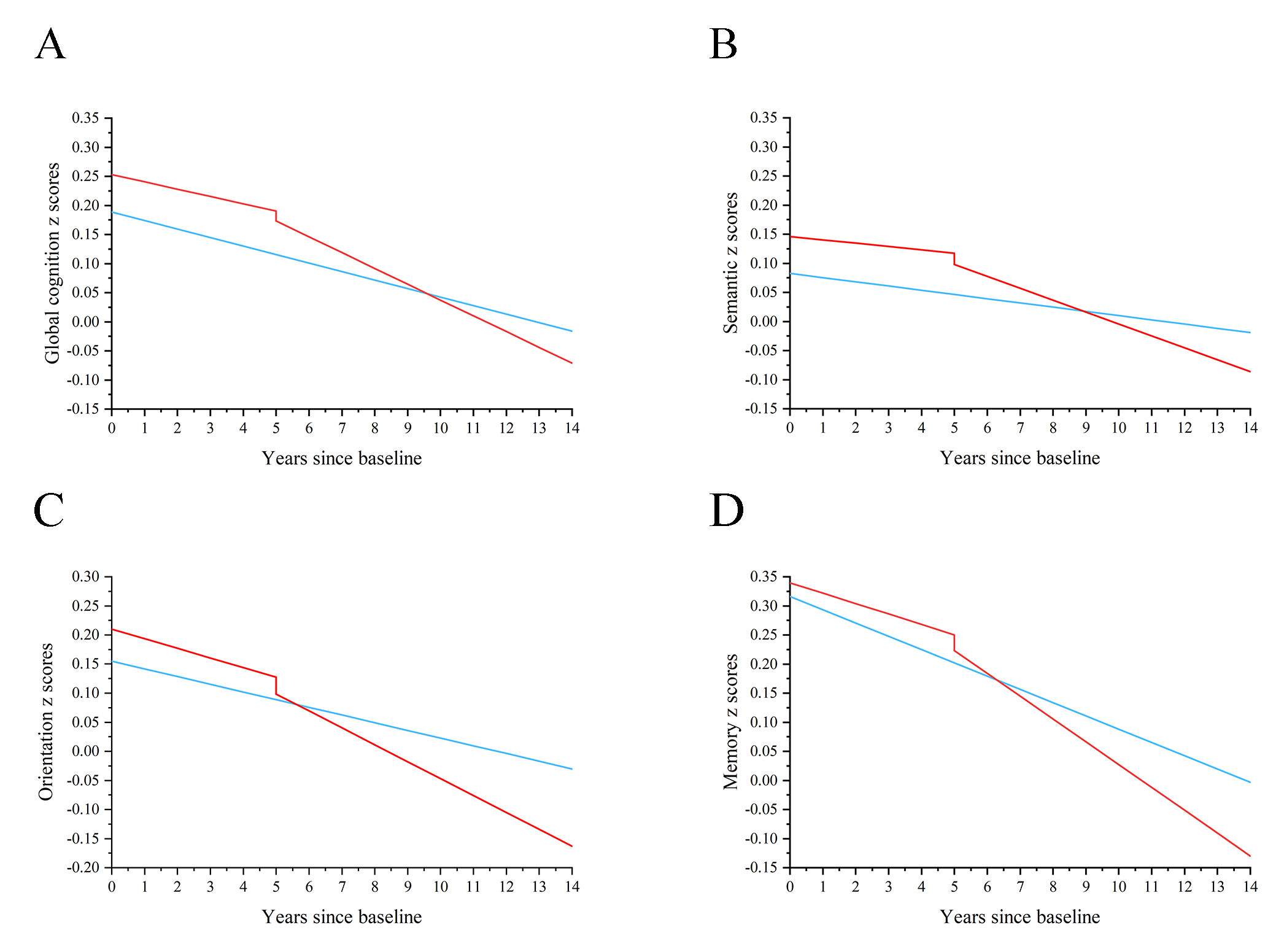

**Supplemental Table 14. Primary analysis only including participants who attended all cognitive tests (743 in the hypertension-free group and the 623 in hypertension group)^a^**

|  | Global cognition |  | Semantic fluency |  | Orientation |  | Memory |  |
| --- | --- | --- | --- | --- | --- | --- | --- | --- |
| Variables^b^ | β (95% CI) | *p* | β (95% CI) | *p* | β (95% CI) | *p* | β (95% CI) | *p* |
| Difference in baseline | -0.008 (-0.100, 0.084) | 0.866 | 0.037 (-0.065, 0.138) | 0.476 | -0.049 (-0.132, 0.034) | 0.248 | -0.092 (-0.179, -0.005) | 0.038 |
| Slope of hypertension-free group | -0.007 (-0.012, -0.003) | <0.001 | -0.002 (-0.007, 0.003) | 0.420 | -0.009 (-0.015, -0.003) | 0.002 | -0.018 (-0.023, -0.014) | <0.001 |
| Difference in slope before hypertension | 0.005 (-0.005, 0.015) | 0.295 | 0.003 (-0.008, 0.014) | 0.583 | 0.005 (-0.007, 0.017) | 0.453 | 0.012 (0.001, 0.022) | 0.026 |
| Changes in slope after hypertension | -0.021 (-0.034, -0.008) | 0.001 | -0.015 (-0.029, -0.001) | 0.031 | -0.010 (-0.025, 0.005) | 0.191 | -0.028 (-0.040, -0.015) | <0.001 |

^a^Adjusted for baseline age, sex, education, marital status, current smoking, current drinking, BMI, physical activity, depression, diabetes, cancer, chronic lung diseases, and heart failure.

^b^Detailed description for variables is shown in Supplemental Figure 1.
